# Gastrointestinal tract symptoms in coronavirus disease 2019: Analysis of clinical symptoms in adult patients

**DOI:** 10.1101/2020.03.23.20040279

**Authors:** Yong Zhang, Zuneng Lu, Bo Wang, Jinxing Cang, Yonggang Ma

## Abstract

**Objective:** To investigate the clinical presentation of coronavirus disease 2019 (COVID-19), particularly the incidence of gastrointestinal tract symptoms.

**Design:** We enrolled adult COVID-19 patients from a mobile cabin hospital in Wuhan with a definitive diagnosis by SARS-CoV-2 nucleic acid testing. Face-to-face interviews were conducted in which the patient selected COVID-19-related symptoms and report the time of onset and duration of symptoms.

**Results:** A total of 212 adults were enrolled in this study, of which 127 (59.9%) were females, mean age was 48.50 ±13.15 (range: 17-79) years, and mean disease course was 26.78±9.16 (3-60) days. Fever and cough were the most common and earliest clinical symptoms of COVID-19.

Diarrhoea occurred in 43.8% (93/212) of patients, of which 86.0% (80/93) had mushy stools. Nausea and vomiting were also common (20.7%). Diarrhoea lasted for 4.00(2.00-8.85) days and mostly occurred 5.00(0.25-11.00) days after the emergence of the first symptoms. Multiple logistic regression analysis found that diarrhoea was significantly correlated with fatigue [OR2.900,95%CI (1.629-5.164), p<0.0001].

**Conclusions:** Gastrointestinal tract symptoms are common in COVID-19 and most occur during the middle stage of the disease and lasts for a short period of time. Clinicians need to pay greater attention to gastrointestinal tract symptoms of COVID-19.

## INTRODUCTION

Coronavirus disease 2019 (COVID-19) has recently become a global pandemic.^1^ Fever and cough are the most common clinical presentations. ^2-3^ Gastrointestinal tract symptoms such as nausea, vomiting, and diarrhoea are often seen in coronavirus infections such as severe acute respiratory syndrome (SARS) and Middle East respiratory syndrome (MERS).^4-5^ Recent studies report that gastrointestinal tract symptoms are rarely observed in COVID-19 patients,^2-3^ observed in about 3.0% of cases. However, a US report revealed that the first patient experienced 2 continuous days of diarrhoea on Day 6 after disease onset.^6^ Are gastrointestinal tract symptoms rare in COVID-19? We recently worked in a mobile cabin hospital and conducted a detailed investigation of the clinical symptoms of patients with COVID-19.

## METHODS

### Ethical considerations

This study was reviewed by the ethics committee of Renmin Hospital of Wuhan University (Ethics Approval No.: WDRY2020-K033) and written informed consent from patients was waived. Only verbal explanation was required, and verbal informed consent was obtained from the patients.

### Subject population

Hardcopy forms were formulated and printed. From February 18 to March 2, 2020, COVID-19 patients ≥16 years of age in Wuchang mobile cabin hospital were recruited. The patients themselves completed a form that collected general demographic information, current medical history, past medical history, date of first positive SARS-CoV-2 nucleic acid result, chest CT result, and COVID-19 contact history. COVID-19-related clinical symptoms were listed with tick boxes in the form. The study participants were asked to check the boxes of their recent symptoms and write down the specific date of onset and duration of each symptom. All patients in this mobile cabin hospital were long-time residents of Wuhan and tested positive for SARS-CoV-2 nucleic acid by reverse transcription polymerase chain reaction (RT-PCR), and their diseases were classified as mild and moderate COVID-19 according to the diagnostic criteria in Diagnosis and Treatment Plan of Coronavirus Disease 2019 (trial edition 5).^7^ After patients completed the hardcopy forms, photographs were taken, and the data were uploaded onto the network. Before the mobile cabin hospital was closed, the hardcopy forms were destroyed as medical waste.

### Data analysis

SPSS 26.0 software was used for statistical analysis. Disease course referred to the duration from onset of first symptoms to the date of the investigation. Disease course in asymptomatic patients was defined as the interval from test date of the positive SARS-CoV-2 nucleic acid test result to the date of the investigation. Symptom duration was defined as the interval from onset of that symptom to resolution or investigation. Symptom occurrence node refers to the duration from onset of the first symptom of COVID-19. High fever was defined as temperature ≥39°C. Patients with ages ≥50 years were described as elderly patients. A disease course ≥20 days was defined as long disease course. A history of chronic disease was defined as other diseases that persisted for 3 months before COVID-19 such as hypertension, diabetes, hepatitis B, and tuberculosis. The Kolmogorov-Smimov test was used to determine whether the data were normally distributed.

Continuous variables were expressed as the means and standard deviations or medians and interquartile ranges (IQR) as appropriate. Categorical variables were summarized as the counts and percentages in each category. We grouped patients into diarrhoea and non-diarrhoea□Wilcoxon rank-sum tests were applied to continuous variables, chi-square tests and Fisher’s exact tests were used for categorical variables as appropriate. To identify risk factors for COVID-19, Pearson’s chi-squared test was used for correlation analysis. Differences with a P-value < 0.05 were considered statistically significant.

## RESULTS

A total of 212 adults were enrolled in the study, of which 127 (59.9%) were females, and mean age was 48.50 ±13.15 (range: 17–79) years. In terms of educational level, 8.0%(17/212) completed elementary school education, 52.8%(112/212) attained middle school education, and 39.2% (83/212) were college graduates. Mean disease course was 26.78 ± 9.16 (range: 3–60) days. There were 12 clinically asymptomatic patients, and mean disease course was 18.08 ± 9.32 (range: 9–33) days. Five of these 12 patients had chest computed tomography (CT) scans that showed viral pneumonia but they had no clinical presentation. Approximately 44.3% of the patients claimed to have a clear contact history with individuals with COVID-19. Chest CT scans showed that almost all patients (96.7%, 207/212) in this study had specific characteristics for COVID-19.^7^

Fever and cough were the most common clinical presentations (Figure 1), and patients with high-grade fever accounted for 14.2% (30/212). Profuse sweating occurred in 114 patients, of which 89.5% (102/114) had night sweats. Diarrhoea occurred in 43.8% (93/212) of patients, of which 86.0% (80/93) had mushy stools. Nausea and vomiting were also common (20.7%). Around 3.3% (7/212) of the patients developed conjunctival congestion.

**Figure 1.**
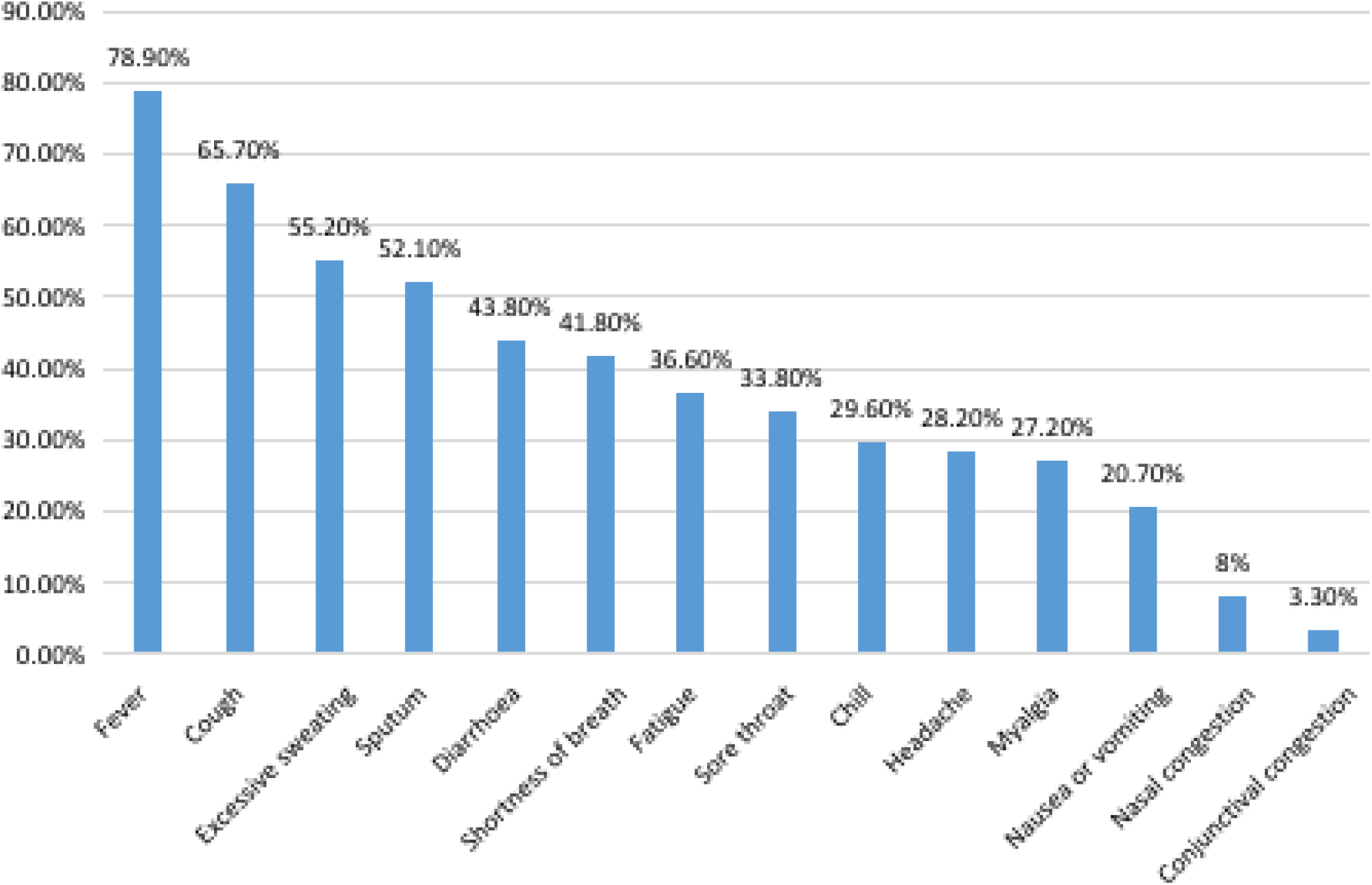
Incidence of clinical symptoms

Cough was the most persistent symptom, whereas conjunctival congestion had the shortest duration. Diarrhoea persisted for an median duration of 4.00 (2.00-8.75) days and is one of the clinical symptoms with the shortest duration.

Nasal congestion, myalgia, headache, chills, fever, fatigue, and coughing were common initial symptoms, whereas gastrointestinal tract symptoms were rare initial symptoms. In this study, 5 (2.4%) patients had an initial symptom of diarrhoea, whereas most patients developed diarrhoea 5.00(0.25-11.00) days (median) after the development of initial symptoms (Figure 3).

**Figure 2.**
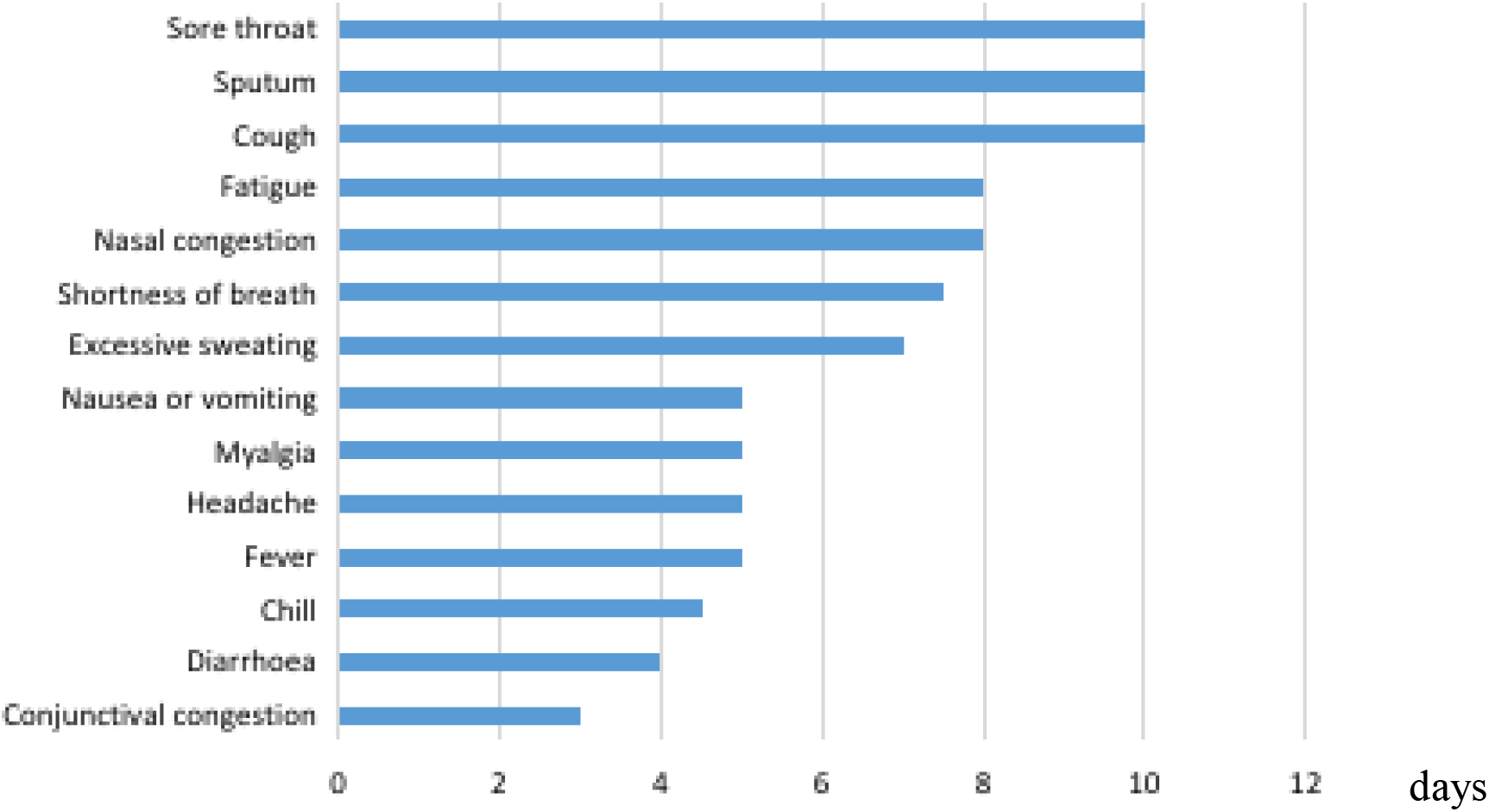
Duration of clinical symptoms Notes: the data above is medium.

**Figure 3.**
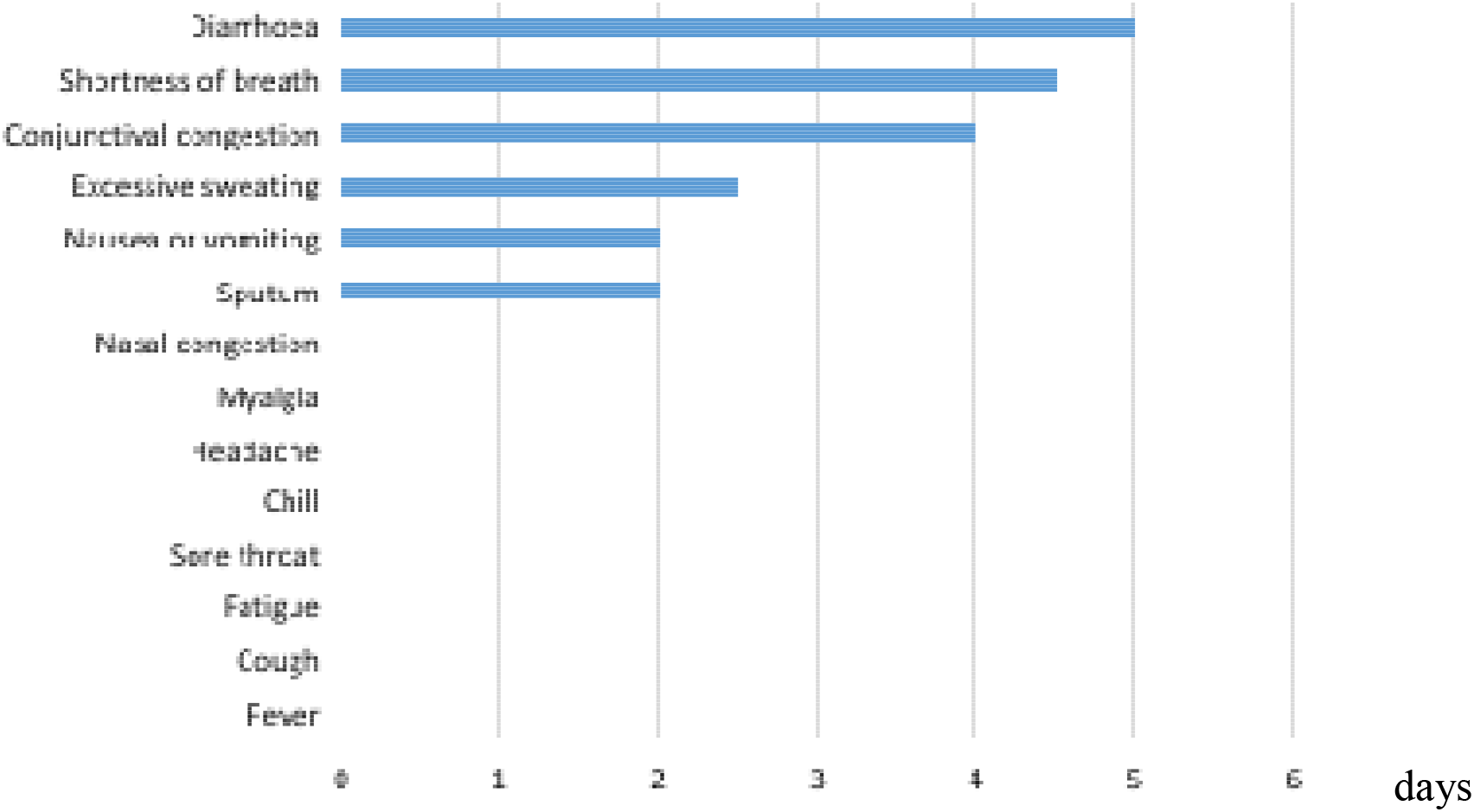
Occurrence node of clinical symptoms Notes: the data above is medium.

Correlation analysis of diarrhoea found that fatigue had the highest correlation with diarrhoea□OR, 2.900, 95%CI(1.629-5.164), p<0.0001□. Diarrhoea was not associated with sex or disease course(P>0.05), and old age might be a protective factor against diarrhoea (OR, 0.552, 95%CI (0.319-0.955))(Table 1).

**Table 1.**
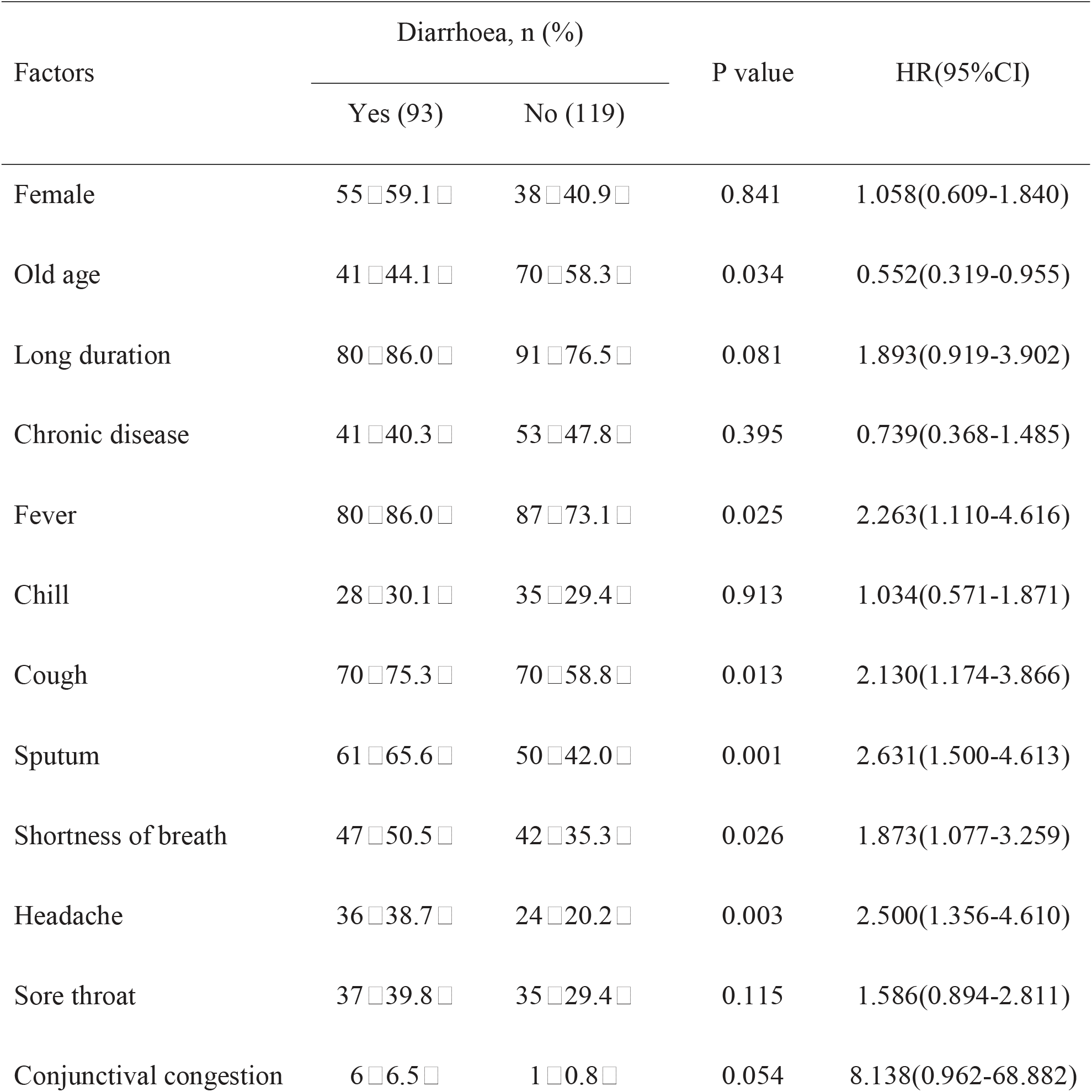
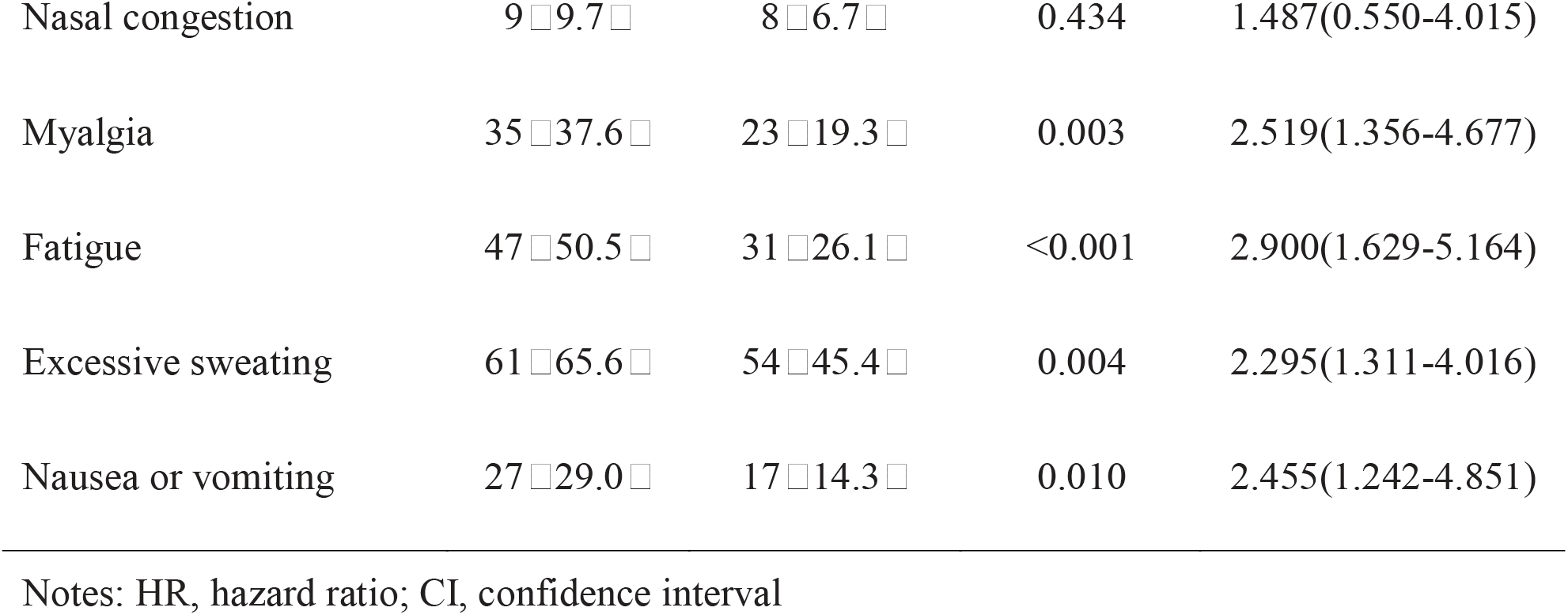
The related factors with diarrhoea

| Factors | Diarrhoea, n (%) |  | P value | HR(95%CI) |
| --- | --- | --- | --- | --- |
|  | Yes (93) | No (119) |  |  |
| Female | 55□59.1□ | 38□40.9□ | 0.841 | 1.058(0.609-1.840) |
| Old age | 41□44.1□ | 70□58.3□ | 0.034 | 0.552(0.319-0.955) |
| Long duration | 80□86.0□ | 91□76.5□ | 0.081 | 1.893(0.919-3.902) |
| Chronic disease | 41□40.3□ | 53□47.8□ | 0.395 | 0.739(0.368-1.485) |
| Fever | 80□86.0□ | 87□73.1□ | 0.025 | 2.263(1.110-4.616) |
| Chill | 28□30.1□ | 35□29.4□ | 0.913 | 1.034(0.571-1.871) |
| Cough | 70□75.3□ | 70□58.8□ | 0.013 | 2.130(1.174-3.866) |
| Sputum | 61□65.6□ | 50□42.0□ | 0.001 | 2.631(1.500-4.613) |
| Shortness of breath | 47□50.5□ | 42□35.3□ | 0.026 | 1.873(1.077-3.259) |
| Headache | 36□38.7□ | 24□20.2□ | 0.003 | 2.500(1.356-4.610) |
| Sore throat | 37□39.8□ | 35□29.4□ | 0.115 | 1.586(0.894-2.811) |
| Conjunctival congestion | 6□6.5□ | 1□0.8□ | 0.054 | 8.138(0.962-68.882) |
| Nasal congestion | 9□9.7□ | 8□6.7□ | 0.434 | 1.487(0.550-4.015) |
| Myalgia | 35□37.6□ | 23□19.3□ | 0.003 | 2.519(1.356-4.677) |
| Fatigue | 47□50.5□ | 31□26.1□ | <0.001 | 2.900(1.629-5.164) |
| Excessive sweating | 61□65.6□ | 54□45.4□ | 0.004 | 2.295(1.311-4.016) |
| Nausea or vomiting | 27□29.0□ | 17□14.3□ | 0.010 | 2.455(1.242-4.851) |

Notes: HR, hazard ratio; CI, confidence interval

## DISCUSSIONS

Coronavirus is a large virus family that can cause various conditions ranging from the common cold to severe infectious diseases such as SARS and MERS.8 COVID-19 is a recently discovered infection in humans and is now considered a pandemic.^1,9^ The most common clinical presentations associated with coronavirus infection are fever and cough.^2-3,9^ The results of this study have further proven previous conclusions. Studies have found that gastrointestinal tractsymptoms are one of the common clinical presentations in SARS and MERS. ^4-5,9^ However, recent studies found that diarrhoea is rare in COVID-19,^2-3,10^ with an incidence of about 3%. A US study reported that their first patient, who had mild COVID-19, developed diarrhoea lasting for 2 days. ^6^ In addition, a study reported that diarrhoea may be an initial symptom of COVID-19.^11^ In this study, we found that a minority of patients had an initial symptom of diarrhoea. In contrast to the results of previous studies, this study found that diarrhoea, nausea, and vomiting are common in COVID-19. Why are there differences between the results of this study and other past clinical investigations? First, the data sources of these studies vary. The information used by Zhong et al. was obtained from medical records, ^3,10^ whereas the data in this study was provided by the patients themselves. Even though this is a retrospective study, the study subjects were adults and most had an educational level of middle school and above, thereby ensuring the reliability and authenticity of the content of this study. Second, the time nodes of the data records are different. The results of this study found that diarrhoea mostly occurred 5 days after the development of initial symptoms; therefore, patients may not have developed gastrointestinal tract symptoms such as diarrhoea, nausea, and vomiting in the other studies. In this study, the mean disease course when patients underwent interviews was 26.78 ± 9.16 days, which far exceeds the duration in other studies. For example, Gong et al. showed that the mean disease course of deceased patients was 10.56 ± 4.42 days.^12^ Furthermore, in contrast to cough and other symptoms, diarrhoea in COVID-19 patients lasted for a short duration and mostly occurred as mushy stools instead of watery diarrhoea in SARS.^4^ Therefore, diarrhoea may be overlooked by clinicians and patients during COVID-19. As digestive tract symptoms such as diarrhoea, nausea, and vomiting tend to be overlooked in coronavirus infections, Kim et al. proposed considering diarrhoea as one of the clinical presentations during MERS diagnosis.^13^

Why do we need to pay attention to gastrointestinal tract symptoms such as diarrhoea, nausea, and vomiting? First, as COVID-19 is a wasting disease, vomiting and diarrhoea affect nutrient absorption in patients and therefore may affect disease outcomes. Our study found that diarrhoea is significantly correlated with fatigue, suggesting that diarrhoea may decrease the quality of life of patients. Hence, clinicians must actively strengthen nutrition supportive treatment in COVID-19 patients. In addition, Xiao et al. isolated viable SARS-CoV-2 virus from fecal samples of COVID-19 patients and the human receptor for SARS-CoV-2, angiotensin-converting enzyme, ^14^ is highly expressed in the stomach, duodenum, and rectal epithelial cells, supporting the possibility that SARS-CoV-2 can enter host cells in the digestive tract. Gu et al. proposed that the digestive tract may be one of the target organs of SARS-CoV-2.^15^ Our study showed that digestive tract symptoms such as diarrhoea, nausea, and vomiting are common in COVID-19, suggesting that the possibility of fecal–oral transmission is high. Therefore, we need to actively promote hand hygiene to prevent this disease. Moreover, public health authorities need to actively manage digestive tract excretions from patients to prevent the spread of COVID-19.

As the subjects of this study are patients with mild cases in mobile cabin hospitals, we were unable to determine whether gastrointestinal tract symptoms are common in severe patients. Therefore, we can only suggest that digestive tract symptoms such as diarrhoea, nausea, and vomiting are common in mild COVID-19. At present, no large-scale study has discovered the SARS-CoV-2 virus in fecal samples. ^14^ Therefore, a large sample size of COVID-19 patients is required for a study in which stool samples should be collected approximately one week after the onset of initial clinical symptoms in COVID-19 patients to assess the proportion of COVID-19 patients with stool samples containing the SARS-CoV-2 virus. This will provide valuable evidence to support aggressive public health strategies.

In conclusion, gastrointestinal tract symptoms are common in COVID-19 and most occur during the middle stage of the disease and lasts for a short period of time. Clinicians need to pay greater attention to the gastrointestinal tract symptoms of COVID-19.

## Data Availability

All data relevant to the study are included in the article or uploaded as supplementary information

## ACKNOWLEDGEMENTS

The authors would like to thank all patients for making this study possible. Mourn to the victims.

## CONTRIBUTORS

YZ conceived and designed the study. BW, JXC and YGM performed the experiments and performed the literature survey. YZ and ZNL contributed to the writing and checking of the letter.

### Funding

No funding.

### Patient consent for publication

Obtained.

### Provenance and peer review

Not commissioned; externally peer reviewed.

### Open access

This is an open access article distributed in accordance with the Creative Commons Attribution Non Commercial (CC BY-NC 4.0) license, which permits others to distribute, remix, adapt, build upon this work non-commercially, and license their derivative works on different terms, provided the original work is properly cited, appropriate credit is given, any changes made indicated, and the use is non-commercial. See: http://creativecommons.org/licenses/by-nc/4.0/.

